# From “Tech Fear” to Digital Self-Efficacy: A COM-B and TDF Analysis of Tele-Exercise Engagement in Older Adults at Risk of Falls

**DOI:** 10.64898/2026.11.15.26367805

**Authors:** Karly Oi-wan Chan, Janet Lok-chun Lee, Ben Yuk-fai Fong, Peter Pok-man Yuen, Fowie Sze-fung Ng, Vincent Tin-sing Law, I-san Cheung, Sam Wing-sum Wong, Tommy Ka-chun Ng, Wilson Chi-pun Fung

## Abstract

**Background:** Tele-exercise offers an alternative approach to physical activity for older adults, however remote delivery introduces distinctive challenges.

**Objectives:** Nested within a 3-month randomised controlled trial (RCT), this qualitative study explored the experiences of community-dwelling older adults at risk of falls who completed synchronous tele-exercise, mapping findings to the COM-B model, Theoretical Domains Framework (TDF), and Behaviour Change Techniques (BCTs) to inform intervention design.

**Methods:** Semi-structured interviews and delivery format preference queries were conducted with 25 purposively sampled older adults following the intervention. Data were analysed using hybrid thematic analysis integrated with COM-B, TDF, and the Behaviour Change Wheel (BCW).

**Results:** Despite baseline digital literacy barriers, participants engaged successfully across six themes: Technological Skills, Access & Support; Coach Support & Exercise Adaptability; Health Motivation & Goal Achievement; Emotional Journey with Technology; Home Environment & Accessibility; and Social Interaction Dynamics. Core BCTs addressing low digital literacy and physical limitations included practical social support (in-home onboarding), environmental restructuring (hardware setup), graded tasks, and social reward. These techniques facilitated an emotional transition from initial apprehension to pride, fostering digital self-efficacy. Notably, an implementation paradox emerged: while over half preferred face-to-face (f2f) delivery, they demonstrated high adherence in tele -exercise programme, indicating that delivery preference does not preclude digital capability.

**Conclusion:** Tele-exercise is feasible and acceptable for older adults at risk of falls, challenging assumptions that low digital literacy precludes digital health adoption. Embedding targeted BCTs within a scaffolded framework addresses structural and psychological barriers, reinforcing digital self-efficacy to sustain remote participation.

**Key Points:**

- Tele-exercise is feasible when behaviour techniques shift participants from digital anxiety to self-efficacy.
- Remote delivery removes transport barriers but requires personalized home-safety checks to address domestic hazards.
- Social connectedness, peer modelling, and instructor feedback transform isolated routines into engaging communal experiences.
- Clinicians should prescribe structured tele-exercise with standardized onboarding and targeted techniques to reduce fall risks.

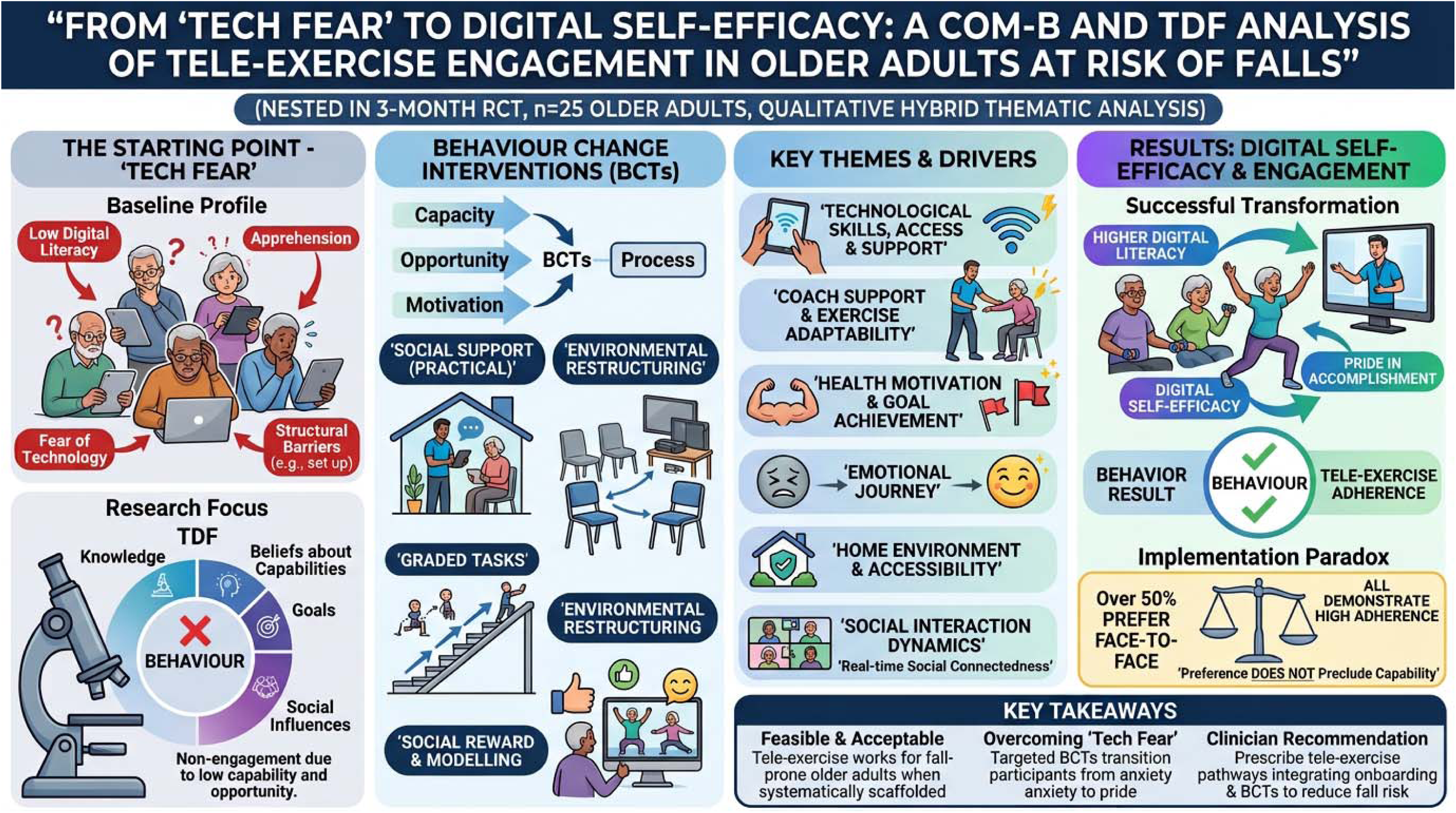

## 1. Introduction

In the past decade, the rapid advancement of information and communications technology (ICT) has made telehealth an important tool for managing health and wellness, especially for those living in remote areas or with limited access to healthcare services. The COVID-19 pandemic has further accelerated its widespread adoption [1]. Telehealth is a broad term that encompasses the remote delivery of health-related services by healthcare professionals. Specifically, tele-exercise refers to the remote delivery of home-based exercise interventions through video-linked appointments using videoconferencing software like Zoom, Skype, FaceTime, or VooV, along with devices such as smartphones, tablets, or computers equipped with webcams and internet connections [2]. The effective adoption of tele-exercise offers significant potential to enhance care continuity, improve individual access, and bypass traditional logistical barriers such as travel and adverse weather, which is particularly beneficial for community-dwelling older adults at risk of falls [3,4].

Recent systematic reviews demonstrate that tele-exercise can achieve clinical outcomes comparable to traditional f2f interventions [5]. Furthermore, qualitative studies exploring the acceptability of recreation- and rehabilitation-oriented tele-exercise reveal that while older adults appreciate the convenience of home delivery, they frequently encounter initial technological barriers [6–8]. Many older adults experience digital exclusion or ’tech fear’, though they can gradually adapt with adequate external support. However, while previous literature successfully describes these subjective barriers, it often fails to systematically translate these lived experiences into actionable, theory-driven strategies for intervention design. Consequently, less attention has been given to the precise mechanisms required to facilitate the behavioural transition from a preference for traditional f2f classes to sustained engagement in a digital modality.

To understand how tele-exercise can be effectively embedded into community care, it is necessary to move beyond simply identifying barriers and toward an in-depth behavioural diagnosis of the individual, social, and environmental factors influencing uptake. Recent research highlights that tailored implementation strategies are crucial for improving the uptake of falls prevention recommendations in practice [9] The Behaviour Change Wheel (BCW) provides a comprehensive framework for this translation [10]. At its core, the Capability, Opportunity, Motivation-Behaviour (COM-B) model acts as a behavioural system, positing that engagement requires adequate Capability, Opportunity, and Motivation. The Theoretical Domains Framework (TDF) expands this by subdividing the components into granular domains, allowing for a precise analysis of the theoretical mechanisms at play [11]. Crucially, these diagnostic domains can then be mapped to the Behaviour Change Technique Taxonomy v1 (BCTTv1), which provides the specific “active ingredients” (e.g., graded tasks, practical social support) needed to directly target identified deficits [10].

Therefore, the aim of this study was to qualitatively explore the experiences and perceptions of community-dwelling older adults at risk of falls who participated in a synchronous tele-exercise programme. By utilising the COM-B model and TDF as theoretical lenses, this study sought to identify the precise determinants and facilitating factors influencing the engagement and uptake of this novel delivery mode. Furthermore, by linking these theoretical domains to specific BCTs, this research aims to provide actionable recommendations for designing future, multi-option tele-exercise frameworks that effectively foster sustained physical activity behaviour change in aging populations.

## 2. Methodology

The study was nested within a RCT comparing synchronous group-based tele-exercise (TE) versus community-based (CB) exercise for older adults at risk of falls. The study was approved by the Ethics Committee of the first author’s institution (RC/ETH/H/0029). The study was registered in the Chinese Clinical Trial Registry (ChiCTR2200063370) on 5 September 2022. The study protocol has been published [12], with quantitative results under review [13]. Preliminary descriptive findings from this dataset were previously presented as a conference abstract [14]. This manuscript reanalyses qualitative data, mapping to COM-B, TDF, and BCTs to inform intervention design.

The intervention was a modified Otago Exercise Programme (OEP) delivered over three months. TE sessions were held via Zoom in participants’ homes, with devices provided as needed. All participants attended thrice-weekly, 45-minute classes (8–11 per group). Sessions followed a structured format—warm-up, OEP [15], “workouts on demand,” cool-down—led by an instructor in their late 60s experienced with teaching older adults. Exercises were tailored (e.g., shorter holds, fewer reps for low strength), with individual feedback during and after sessions [12].

### Study design

This qualitative study used semi-structured interviews to explore barriers and enablers of tele-exercise training in older adults at risk of falls. Analysis was guided by the TDF and the COM-B model. The TDF consolidates constructs from multiple behaviour change theories into 14 domains (e.g. knowledge, skills, role, self-efficacy, environment) [16], mapped onto COM-B’s components: Capability, Opportunity, and Motivation [17].

Thematic analysis followed Braun & Clarke’s 15-point checklist [18] and six-step process [19], from transcription to reporting, to identify key themes.

### Sampling and recruitment

Participants were recruited from the tele-exercise arm of the RCT [13]. Eligible participants were community-dwelling adults aged 65 or above, at risk of falls, Cantonese-speaking, able to walk unaided, and cognitively fit (MMSE >25); each received a local supermarket voucher worth approximately USD $13 equivalent after completing the interview.

Purposive sampling maximised variation in age, attendance, education, and technology access. Demographic and engagement data are shown in Tables 1–2.

**Table 1.** Demographic and Programme Engagement Characteristics of Participants.

| No. of participants (n) | 25 |
|---|---|
| Mean Age (range) | 76.1±6.5 (65-91) |
| Sex distribution<br>% of male<br>% of female | 20%<br>80% |
| Education level %<br>1. Junior primary or below<br>2. Senior primary<br>3. Junior Secondary<br>4. Senior Secondary<br>5. University or above | 16%<br>36%<br>12%<br>32%<br>4% |
| % Home wi-fi access | 40% |
| % smartphone access | 92% |
| Solitude status | 52% |
| Mean attendance rate (%) (range) | 85.5±10.9% (55.6-100%) |
| PACES score (range) | 108.7±8.3 (95-121) |

\*PACES: Physical Activity Enjoyment Scale, max score:126, with higher scores reflecting higher levels of enjoyment [25].

**Table 2.** Individual Demographic, Technological Access, and Programme Engagement Characteristics of Participants.

| ID | Age range | Solitude status (Y/N) | Frequency of home visit during intervention | Sex | Education level | Home Wi-Fi access (Y/N) | Smartphone access (Y/N) | Adherence rate (%) | PACES score | Prior experience of tele-exercise (Y/N) |
|---|---|---|---|---|---|---|---|---|---|---|
| P1 | 90–94 | N | 8 | F | 1 | N | N | 97.20% | 100 | Y |
| P2 | 70–74 | N | 3 | M | 5 | Y | Y | 91.70% | 120 | N |
| P3 | 75–79 | Y | 6 | M | 2 | N | Y | 94.40% | 116 | N |
| P4 | 75–79 | N | 6 | F | 2 | Y | Y | 83.30% | 101 | N |
| P5 | 80–84 | Y | 8 | F | 1 | N | Y | 66.70% | 95 | N |
| P6 | 65–69 | N | 5 | F | 2 | N | Y | 100.00% | 119 | Y |
| P7 | 75–79 | N | 5 | F | 3 | N | Y | 80.60% | 120 | N |
| P8 | 75–79 | Y | 7 | F | 4 | N | Y | 86.10% | 120 | N |
| P9 | 70–74 | N | 2 | M | 4 | Y | Y | 55.60% | 101 | N |
| P10 | 65–69 | N | 2 | F | 2 | N | Y | 86.10% | 102 | N |
| P11 | 85–89 | Y | 5 | F | 2 | N | Y | 80.60% | 110 | N |
| P12 | 75–79 | Y | 7 | F | 2 | N | Y | 80.60% | 103 | N |
| P13 | 85–89 | Y | 8 | F | 1 | N | N | 69.40% | 104 | N |
| P14 | 70–74 | Y | 1 | F | 4 | Y | Y | 94.40% | 102 | N |
| P15 | 80–84 | N | 5 | M | 2 | N | Y | 83.30% | 102 | N |
| P16 | 65–69 | N | 2 | F | 2 | Y | Y | 94.40% | 96 | N |
| P17 | 75–79 | Y | 4 | F | 4 | Y | Y | 94.40% | 121 | N |
| P18 | 75–79 | N | 7 | F | 4 | Y | Y | 86.10% | 106 | N |
| P19 | 75–79 | N | 4 | F | 3 | Y | Y | 94.40% | 105 | N |
| P20 | 70–74 | Y | 6 | F | 2 | Y | Y | 91.70% | 112 | Y |
| P21 | 70–74 | Y | 6 | F | 4 | Y | Y | 83.30% | 109 | Y |
| P22 | 75–79 | Y | 6 | F | 3 | N | Y | 94.40% | 113 | N |
| P23 | 80–84 | N | 12 | M | 1 | N | Y | 97.20% | 114 | N |
| P24 | 70–74 | Y | 12 | F | 4 | N | Y | 80.60% | 120 | N |
| P25 | 65–69 | Y | 11 | F | 4 | N | Y | 72.20% | 106 | N |

\*Education level: 1: Junior primary or below; 2: senior primary; 3: Junior secondary; 4: Senior secondary; 5: University or above

All participants completed interviews after the 3-month intervention, with recruitment ending once variation targets were met and data saturation confirmed [20]—no new themes emerged between the 23rd and 25th interviews.

### Interview process

Written informed consent was obtained before semi-structured interviews, conducted March–July 2023 either f2f in community centres or via Zoom, depending on preference. All sessions were held privately to ensure confidentiality.

Interviews were conducted by the researcher 1 (MSc, male) and researcher 2 (PhD, female), trained by researcher 3 (PhD, female), an experienced qualitative researcher. No prior relationship existed; participants only knew the team as academic investigators delivering and evaluating the tele-exercise programme.

A semi-structured guide, based on TDF domains and literature [21,22], was pilot-tested with four participants (supplementary material S1). As only minor probes were added, these pilots were included in analysis. Researcher 1 then served as the primary interviewer.

All interviews were video-recorded with consent, averaged 32 minutes (range 23–50), and no repeat interviews were conducted. The COREQ 32-item checklist [23] is provided as supplementary material (S2).

### Data analysis

#### Qualitative Thematic Analysis and Theoretical Diagnosis

All interviews were transcribed verbatim by a third party, with participants anonymized and labelled numerically (#1–25). Data were managed using NVivo 14 (QSR International, Cambridge, Massachusetts, USA) and were analysed using a hybrid deductive-inductive approach strictly aligned with [18] six-phase thematic analysis, integrated with the COM-B and TDF procedures described by [16].

1. **Familiarisation:** Researchers repeatedly read transcripts to gain contextual understanding of participants’ experiences.
2. **Initial coding:** Two researchers (researcher 2 and 3) deductively coded data to TDF domains and inductively captured barriers/enablers. Raw agreement was 79%, used diagnostically to refine coding.
3. **Theme search:** Inductive codes were grouped into sub-themes within COM-B/TDF categories.
4. **Theme review:** Themes were cross-checked against data; discrepancies resolved through consensus until 100% agreement.
5. **Defining themes:** Each theme was refined and defined; remaining transcripts coded using the finalized codebook (S3) by the researcher 1 and 2.
6. **Synthesis:** Themes and domains were integrated into a behavioural diagnosis matrix (Table 3) with illustrative quotations.

**Table 3.** Behavioural Diagnosis of Tele-Exercise Engagement.

| Theme | Subtheme (Barrier / Facilitator) | COM-B Component(s) | TDF Domain(s) | Illustrative Participant Quote |
|---|---|---|---|---|
| Theme 1: Technological Skills, Access & Support | Barrier: Technological Limitations Affecting Participation | • Capability - Psychological<br>• Opportunity - Physical | • Skills<br>• Emotion<br>• Environmental context and resources | *“The screen is too small. When we're doing the exercises, it's hard to see the coach's little movements. So you have to really pay attention, like when he raises his heel a little bit.”*<br>(P3) |
|  | Facilitator: Confidence through Guided Practice | • Capability - Psychological<br>• Opportunity - Social<br>• Motivation - Reflective | • Skills<br>• Beliefs about capabilities<br>• Emotion | *“The difficulty was at the beginning, not knowing how to operate the equipment. You guys (research team) showed me how to do it. It's much better now. At first, I didn't know how to turn the computer tablet on, but you taught us, and now we know.”*<br>(P8) |
| Theme 2: Coach Support & Exercise Adaptability | Barrier: Limited Responsiveness in Online Supervision | • Opportunity - Social<br>• Motivation - Reflective | • Environmental context and resources<br>• Social influences<br>• Skills | *“(Talking to the coach, you wonder if he can hear you) I've never said anything before, because he's never called my name.”*<br>(P5) |
|  | Facilitator: Supportive Coaching for Effective Participation | • Opportunity - Social<br>• Motivation - Reflective<br>• Motivation - Automatic | • Social influences<br>• Beliefs about capabilities<br>• Emotion | *“If I can't do something, the coach will help me during the lesson. Like if I can't lift my arms, he'll tell me to use my hand to tap my shoulder (an easier version) so we can hear each other. He taught me ways to improve my balance and endurance. Now I feel confident I can prevent falls.”*<br>(P1) |
| Theme 3: Health Motivation & Goal Achievement | Barrier: Movement Challenges Linked to Health Conditions | • Capability - Physical | • Skills | *“Balance is the hardest thing, especially with my feet. I have some issues with my left foot, but everything else is fine. It's just the left foot. Some movements are just too hard for me, they're really hard. My balance is really bad, so those movements are difficult.”*<br>(P24) |
|  | Facilitator: Multidimensional Health Gains and Habitual Routine Fostering Functional Independence | • Capability - Physical<br>• Motivation - Reflective<br>• Motivation - Automatic | • Beliefs about capabilities<br>• Beliefs about consequences<br>• Reinforcement<br>• Behavioural regulation | *“I hope I can do more exercise. I think I'll be better at balancing, and my knees will be stronger. I can tell my thigh muscles are a lot more toned now. That gives me more confidence going downstairs, so I'm not scared of tripping forward anymore. It's a huge relief, and I'm getting more exercise too.”*<br>(P3) |
| Theme 4: Emotional Journey with Technology | Barrier: Initial Fear and Anxiety | • Motivation - Reflective<br>• Opportunity - Social | • Beliefs about capabilities<br>• Emotion | *“I don’t know how to press it, so I’ll just let it be. There's nothing else you can do; you just have to sit there and wait.”*<br>(P13) |
|  | Facilitator: Pride and Happiness After Mastering Technology | • Motivation - Reflective<br>• Motivation - Automatic | • Beliefs about capabilities<br>• Emotion<br>• Social influences | *“I feel happier. It’s like my hands and feet are more daring now. There are many things to learn, and even if you don’t fully grasp everything, you still benefit from it. I told my son 'Wow, your mom is really impressive now! Look how smart I am!’ My son replied ‘You can handle a computer tablet even though you can’t use a phone properly yet.’”*<br>(P4) |
| Theme 5: Home Environment & Accessibility | Barrier: Constraints of the Home Environment | • Opportunity - Physical<br>• Opportunity - Social | • Environmental context and resources<br>• Social influences | *“There are many things at home, and my family buys a lot of stuff. There's little space, and the bed is filled with food.”*<br>(P1) |
|  | Facilitator: Convenience of Exercising Without Travel | • Opportunity - Physical<br>• Capability - Physical | • Environmental context and resources | *“When I go to the centre, I have to rush there half an hour early. But here, I just turn on the tablet first — then I can do whatever I want, and when it’s time, I just step over and join in. Hahaha!”*<br>(P5) |
| Theme 6: Social Interaction Dynamics | Barrier: Limited Real-time Chat & Domestic Distractions | • Opportunity - Social | • Environmental context and resources<br>• Social influences | *“(Have you ever chatted with other students during class?) No, never......It's class, and that's the coach! It's disrespectful to chat while the coach is teaching. You can't just pretend the coach isn't there!”*<br>(P25) |
|  | Facilitator: Dual Mechanisms: Offline Community Bonding vs. In-Class Cognitive Motor Focus | • Opportunity - Social<br>• Capability - Psychological | • Reinforcement<br>• Memory, attention and decision processes | *“I am happier now because I have learned something, and we get to interact with each other (after class in the community centre).”*<br>(P4)<br>*“(Socializing is) Not too important, since it's just exercise... But I'm worried that after you get familiar with them, they might come over and chat with you for a long time... I really like to focus.”*<br>(P22) |

#### Behavioural Translation, BCT Mapping, and Intervention Recommendations

To translate the qualitative empirical diagnosis into actionable intervention strategies, an auditable four-step translation procedure was undertaken following the BCW guide [17] and COREQ reporting standards:

1. **Select intervention functions:** Using COM-B/TDF diagnostics (Table 3), functions such as Training, Environmental restructuring, and Enablement were identified to address key barriers and facilitators.
2. **Map BCTs:** Behaviour Change Techniques from BCTTv1 [24] were chosen to operationalise each function.
3. **Appraise recommendations:** Proposed modifications were evaluated against APEASE criteria (Acceptability, Practicability, Effectiveness, Affordability, Safety, Equity) [10].
4. **Feasibility review:** The mapping matrix of RCT components and recommendations was finalized through iterative author consensus, with input from the delivery team to ensure feasibility, safety, and fidelity.

#### Descriptive Analysis of Delivery Format Preferences

Within the Social Influences domain, participants were asked their delivery format preference (f2f, online, or none). Responses were extracted, categorised, and descriptively analysed using frequency counts and percentages to complement the qualitative findings.

## Results

### Participants characteristics

All 25 invited participants consented to join the study. They were older adults (mean age□=□76.1, SD□=□6.5; range 65–91), predominantly female (80%), with varied education—most completed senior primary (36%), while only 4% held university degrees. Technology access was moderate: 92% owned smartphones, but only 40% had home Wi-Fi. Programme adherence was high (mean attendance□=□85.5%, SD□=□10.9; range□55.6–100), and enjoyment was strong (mean PACES□=□108.7, SD□=□8.3; range□95–121).

### Themes Emerging From COM-B and TDF Mapping of Tele-Exercise Experiences

Six themes were identified to describe the tele-exercise experiences of older adults at risk of falls and to identify the behavioural mechanisms influencing their engagement. These themes were: (1) technological skills, access, and support; (2) coach support and exercise adaptability; (3) health motivation and goal achievement; (4) emotional journey with technology; (5) home environment and accessibility; and (6) social interaction dynamics. These themes and their behavioural mechanisms were mapped onto the COM-B model and TDF domains (Table 3) and operationalised into BCW intervention functions, active BCTTv1 ingredients, and future recommendations (Fig 1, Supplementary material S4).

**Figure 1.**
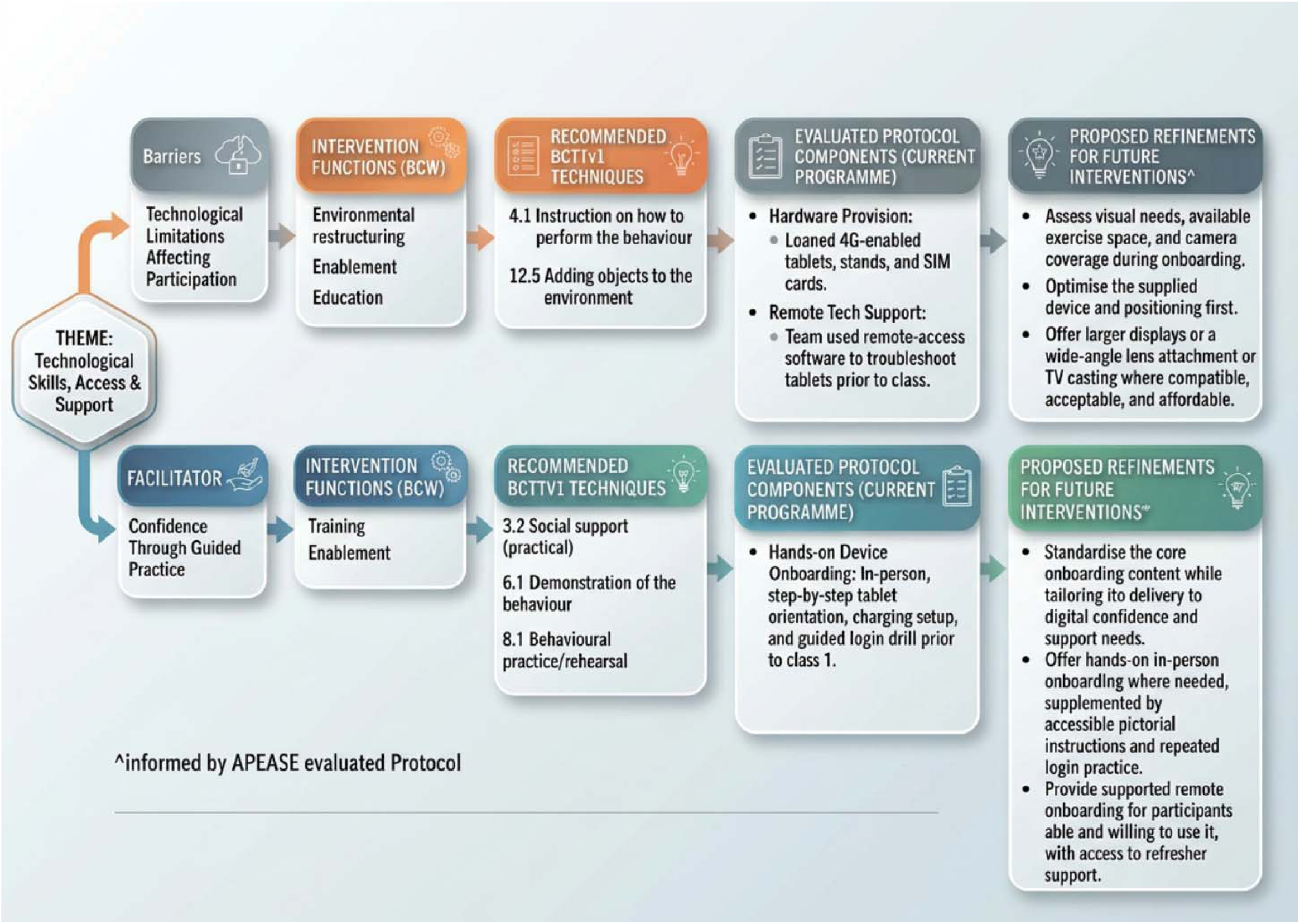
Example mapping of barriers and facilitators to BCW intervention functions and BCTTv1 techniques, with proposed refinements appraised against APEASE.

### Theme #1:Technological Skills, Access and Support

#### Sub-theme: Barriers - Technological Limitations Affecting Participation

For a cohort where 52% had primary education or below, unfamiliarity with digital devices represented an initial deficit in Psychological Capability (COM-B). P1 (age 90-94, Edu 1, attendance 97.2%, PACES 100) noted that the fear of making irreversible errors induced immediate apprehension: “*At first, it might be intimidating if you don’t understand, but once you do, there’s no need to worry. If you don’t know how, you might mess things up.*”

This hesitation acted as a cognitive gatekeeper before physical exercise began. Lacking environmental scaffolding (COM-B: Physical Opportunity), minor glitches provoked acute helplessness, enforcing dependency on younger relatives: “*We older folks tend to be a bit scared of new technology. If something goes wrong, we don’t know how to fix it, so we have to ask younger people for help*” (P9, age 70-74, Edu 4, attendance 55.6%, PACES 101).

Hardware and connectivity constraints further restricted Physical Opportunity (COM-B). P2 (age 70-74, Edu 5, attendance 91.7%, PACES 120) described environmental instability in rural village housing, where poor broadband weakened reception and disrupted exercise routines. Crucially, small tablet displays created safety-critical barriers to motor skill acquisition (TDF: Skills): *“The screen is too small. When we’re doing the exercises, it’s hard to see the coach’s little movements… like when he raises his heel a little bit”* (P3, age 75-79, Edu 2, attendance 94.4%, PACES 116). Inability to discern subtle postural adjustments heightened fall anxiety during unspotted home training.

#### Sub-theme: Facilitators - Confidence Through Guided Practice

Hands-on onboarding by the research team provided vital cognitive scaffolding, restoring Psychological Capability (COM-B) through step-by-step demonstration (BCT 6.1), guided rehearsal (BCT 8.1), and practical social support (BCT 3.2). P5 (age 80-84, Edu 1, attendance 66.7%, PACES 95) shared: *“I know how to turn on the computer tablet, but I have trouble with the devices. But you guys taught me how to adjust the volume on the speakers, right? And turning on… and using the tablet are pretty much the same as turning it off.”* Mastering basic operations generated an immediate surge in Beliefs about Capabilities (TDF): *“The difficulty was at the beginning… You guys showed me how to do it. It’s much better now”* (P8, age 75-79, Edu 4, attendance 86.1%, PACES 120).

Rapid technical troubleshooting functioned as essential Social Opportunity (COM-B), mitigating transient tech-anxiety (TDF: Emotion). Support ranged from optimizing audio amplification (P1) to resolving live connection failures: *“Even if sometimes I feel anxious and can’t log into the class… you quickly found a reason for it… addressing the problems right away”* (P18, age 75-79, Edu 4, attendance 86.1%, PACES 106). Prompt resolution absorbed the emotional burden of technical failure, allowing participants to reserve cognitive capacity for physical training.

### Theme #2:Coach Support & Exercise Adaptability

#### Sub-theme: Barriers - Limited Responsiveness in Online Supervision

The virtual interface diminished the immediacy of f2f interaction, creating safety concerns and weakening behavioural reinforcement. P5 described feeling invisible: *“(Talking to the coach, you wonder if he can hear you) I’ve never said anything before, because he’s never called my name.”* This perceived absence of verbal acknowledgment undermined Social Opportunity (COM-B) and Reflective Motivation, rendering participants passive viewers. Furthermore, latency in error correction disrupted real-time feedback loops essential for safe balance acquisition (TDF: Skills, Environmental Context and Resources): *“In a physical class, the instructor would respond immediately. However, in a computer-based environment, the response wouldn’t be instantaneous”* (P12, age 75-79, Edu 2, attendance 80.6%, PACES 103).

#### Sub-theme: Facilitators - Supportive Coaching for Effective Participation

The coach’s adaptive instruction bridged the physical distance, serving as the primary catalyst for self-efficacy. By offering regressed movements, coaches utilized Graded Tasks (BCT 8.7) to prevent failure and foster mastery (TDF: Beliefs about Capabilities): *“If I can’t lift my arms, he’ll tell me to use my hand to tap my shoulder… He taught me ways to improve my balance and endurance. Now I feel confident I can prevent falls”* (P1).

Synchronous coaching provided continuous visual modelling (TDF: Social Influences), stimulating Automatic and Reflective Motivation (COM-B): *“the coach explains everything so clearly. When he demonstrates, it’s really clear… The coach does it with you, so it motivates you to do it”* (P3). P2 highlighted the value of percentage-based effort scaling: *“The coach also adjusts the exercises… ’Just do 70% or 60% of it, that’s enough.’ I was a bit nervous at first, but I realized later that this course is pretty suitable and… gradual.”* Scaling exertion to 60–70% capacity (BCT 8.7) provided psychological safety, downregulating negative affective arousal (TDF: Emotion) and turning performance apprehension into attainable routine (TDF: Beliefs about Capabilities).

### Theme #3: Health Motivation & Goal Achievement

#### Sub-theme: Barriers - Movement Challenges Linked to Health Conditions

Pre-existing musculoskeletal impairments exposed baseline deficits in Physical Capability (COM-B) and motor stamina (TDF: Skills). P9 noted that repeated unilateral movements aggravated left foot weakness (*“repeat the same movement three or four times… makes it a bit harder”*), while P12 admitted: *“I stop when he stops. But it’s really tough. I mean, my legs just aren’t strong enough.”* P24 (age 70-74, Edu 4, attendance 80.6%, PACES 120) explicitly linked balance deficits to fall-related fear: *“Balance is the hardest thing… Some movements are just too hard for me… My balance is really bad.”* These deficits emphasise that balance interventions in frail cohorts require carefully curated biomechanical loads to prevent injury and disengagement.

#### Sub-theme: Facilitators - Multidimensional Health Gains and Habitual Routine Fostering Functional Independence

Tangible physiological improvements acted as powerful operant conditioning (TDF: Reinforcement), strengthening Reflective Motivation (COM-B). Enhanced muscle tone directly reduced kinesiophobia: *“I can tell my thigh muscles are a lot more toned now. That gives me more confidence going downstairs, so I’m not scared of tripping forward anymore”* (P3). For P22 (age 75-79, Edu 3, attendance 94.4%, PACES 113), relief from chronic back pain served as a somatic reward, transforming exercise from a chore into a daily self-management ritual.

Participants also reported cognitive restructuring regarding aging: *“Whether it’s thinking, memory, or bodily functions, I feel healthier, and the decline seems to slow down… exercising is very positive for me”* (P3). Functional recovery fostered behavioural spillover into recreational life: *“My legs are stronger now! I went to play ping pong… my legs didn’t feel as tired… I even walk faster now”* (P16, age 65-69, Edu 2, attendance 94.4%, PACES 96). Finally, predictable scheduling (Tuesdays, Thursdays, Saturdays at 11:00 AM; P2, P3) reduced cognitive load, acting as an environmental prompt (TDF: Behavioural Regulation) that converted effortful intention into Automatic Motivation (COM-B) driven by habit.

### Theme #4: Emotional Journey with Technology

#### Sub-theme: Barriers - Initial Fear and Anxiety

Unfamiliarity with technology initially triggered passive avoidance and learned helplessness, reflecting a collapse of Reflective Motivation (COM-B) and low Beliefs about Capabilities (TDF): *“I don’t know how to press it, so I’ll just let it be. There’s nothing else you can do; you just have to sit there and wait”* (P13, age 85-89, Edu 1, attendance 69.4%, PACES 104). P18 voiced acute anxiety regarding compromised autonomy and family reliance (COM-B: Social Opportunity): *“My biggest worry is that if something unexpected happens during class, I won’t know which button to press. My daughter might not be there to help.”*

#### Sub-theme: Facilitators - Pride and happiness after mastering technology

Digital mastery catalysed broader emotional empowerment and intergenerational pride (TDF: Beliefs about Capabilities, Social Influences). P4 (age 75-79, Edu 2, attendance 83.3%, PACES 101) described how technological competence directly altered physical self-perception: *“I feel happier. It’s like my hands and feet are more daring now… I told my son ’Wow, your mom is really impressive now!…’ My son replied ‘You can handle a computer tablet even though you can’t use a phone properly yet.’”* Affective pride (TDF: Emotion) overrode baseline fear of movement. Similarly, P18 transitioned from apprehension to intrinsic anticipation: *“All this information technology stuff has made things easier for us old folks… I’m happy and always looking forward to joining the class.”*

### Theme #5: Home Environment & Accessibility

#### Sub-theme: Barriers - Constraints of the home environment

Domestic settings introduced acute deficits in Environmental Context and Resources (TDF). Exercising in congested living quarters restricted range of motion and elevated collision hazards: *“There are many things at home, and my family buys a lot of stuff. There’s little space, and the bed is filled with food”* (P1). Conflicting family routines introduced competing social demands (COM-B: Social Opportunity; P2), while afternoon sunlight glare disrupted visual tracking (TDF: Environmental Context and Resources): *“There’s glare — the sun from the west causes a lot of reflection”* (P20, age 70-74, Edu 2, attendance 91.7%, PACES 112), forcing participants to guess postures and introducing balance risks.

#### Sub-theme: Facilitators - Convenience of exercising without travel

Eliminating transit burdens conserved limited Physical Capability (COM-B), especially for seniors requiring walking aids: *“I don’t want to go to the centre’s exercise class every day. I have to walk on a slope and use my cane”* (P1). Removing the logistical exhaustion of commuting lowered behavioural friction: *“When I go to the centre, I have to rush there half an hour early. But here, I just turn on the tablet first… and when it’s time, I just step over and join in”* (P5). This frictionless transition anchored the routine within the home environment (TDF: Environmental Context and Resources; COM-B: Physical Opportunity).

### Theme #6: Social Interaction Dynamics

#### Sub-theme: Barriers - Limited Real-Time Chat & Domestic Distractions

Domestic environments blurred classroom boundaries; ambient noise and background televisions from unmuted homes disrupted group focus (COM-B: Social Opportunity; P9). Furthermore, internalized cultural norms of deference toward the instructor suppressed spontaneous horizontal peer communication (TDF: Social Influences): *“It’s class, and that’s the coach! It’s disrespectful to chat while the coach is teaching. You can’t just pretend the coach isn’t there!”* (P25, age 65-69, Edu 4, attendance 72.2%, PACES 106).

#### Sub-theme: Facilitators - Dual Mechanisms: Offline Community Bonding vs. In-Class Cognitive Motor Focus

Social dynamics diverged into two distinct mechanisms:

##### Offline Community Re-anchoring

Rather than forcing online socialization, tele-exercise served as a common ground for peer interaction deferred to community centre visits (COM-B: Social Opportunity; TDF: Reinforcement; P4). P4 shared that ““I am happier now because I have learned something, and we get to interact with each other (after class in the community centre).”

##### Preserving Cognitive-Motor Focus

Other participants actively preferred virtual separation to eliminate social distractions and preserve cognitive bandwidth (TDF: Memory, Attention, and Decision Processes): *“Once the online exercise class is over, that’s it, but communication… isn’t that important”* (P17, age 75-79, Edu 4, attendance 94.4%, PACES 121). P22 (age 75-79, Edu 3, attendance 94.4%, PACES 113) emphasised that avoiding conversational obligations was necessary to maintain balance: *“I’m worried that after you get familiar with them, they might come over and chat… They’re not exercising, just talking… I really like to focus.”*

### Preference on Mode of Exercise Delivery

Preferences for exercise training format were coded into three categories (f2f, online, no preference) and analysed quantitatively using frequency counts.

#### Face-to-Face Preference (56%, n = 14)

The majority preferred in-person classes as a psychological safety net. Immediate error correction transferred the burden of safety back to the expert (TDF: Reinforcement, Skills): *“If I make a mistake, the coach will know right away and correct us immediately”* (P6, age 65-69, Edu 2, attendance 100%, PACES 119). Multi-sensory instruction bridged educational gaps: *“You can actually touch and feel things, which is clearer than online”* (P8). Additionally, physical attendance provided essential Social Opportunity (COM-B) and emotional co-regulation (TDF: Emotion), counteracting chronic loneliness among solitary seniors: *“Having someone to talk to is important. It allows everyone to check in on each other and feel less lonely”* (P9; supported by P15).

#### Online Preference (16%, n = 4)

A focused minority favoured remote classes for visual and cognitive efficiency. Digital camera zoom provided clearer instructional demonstrations than crowded community halls (TDF: Social Influences): *“Because there are fewer people, I can get a close-up view of what the instructor is doing”* (P22). Minimizing social chatter lowered cognitive dual-task demands, allowing undivided attention on balance execution (TDF: Attention, Decision Making, and Memory): *“In a real-life class, there are too many people talking, and it’s harder to focus”* (P23, age 80-84, Edu 1, attendance 97.2%, PACES 114). P24 highlighted personal autonomy: *“Once you log off, everyone can go do their own things… online feels like it’s just me and the coach”* (TDF: Behavioural Regulation).

#### No Clear Preference (28%, n = 7)

Over a quarter expressed situational ambivalence, treating the modalities as complementary:

- **Environmental & Weather Resilience:** Home sessions protected routine adherence during adverse weather (TDF: Environmental Context and Resources): *“Even if it’s freezing cold or raining, you can still exercise at home”* (P2).
- **Physical Energy Conservation:** Travel distance determined modality choice (COM-B: Physical Opportunity): *“If the centre is close by, I’ll go. But if it’s far, I’d rather stay home”* (P11, age 85-89, Edu 2, attendance 80.6%, PACES 110).
- **Perceived Instructional Equivalence:** Highly adapted participants experienced no communication deficits: *“You can call out to him, and they’ll hear you right away, just like f2f”* (P10, age 65-69, Edu 2, attendance 86.1%, PACES 102).

## Discussion

This study applied the COM□B model and TDF to examine older adults’ engagement in synchronous tele□exercise. Key determinants were identified among those facing a “double barrier”—risk of falls combined with lower socioeconomic status and limited technology access. By mapping sub themes to theoretical domains and specific BCTs, the findings yield actionable strategies for designing tele□exercise programmes that can be embedded within community care systems.

### Capability

Technical Errors and Fall Anxiety (COM B: Psychological & Physical; TDF: Skills, Emotion, Beliefs about Capabilities)

In prior literature, technical issues are framed as operational hurdles [26]. For older adults at high fall risk living alone, however, these challenges became safety-critical. With 52% having only primary education, navigation errors often led to acute helplessness (P13; Theme 4). Lacking co-residing family, participants feared misinterpreting cues or audio lags during balance drills could trigger unmonitored falls, undermining Psychological Capability.

Anxiety was mitigated not by practice alone but through coach-mediated adaptability (Theme□2). Calibrated exertion targets (e.g., 60–70% effort, seated variations; P2) provided mastery experiences and verbal persuasion, restoring Beliefs about Capabilities. This structured support offered psychological safety while maintaining physical challenge, consistent with Self-Efficacy Theory [27]. Ultimately, technophobia shifted into pride and exercise self-efficacy (Themes□3 &□4).

### Opportunity

Spatial and Social Trade Offs (COM□B: Physical & Social; TDF: Environmental Context, Social Influences)

The domestic environment functioned as a dual-edged physical opportunity (Theme 5). Remote delivery reduced barriers such as transport and adverse weather [28], especially for mobility-compromised seniors navigating slopes and stairs (P1). Yet, domestic spaces introduced risks—clutter, poor lighting, and limited camera visibility—that compromised exercise execution unless proactively managed [29].

Social opportunity (Theme 6) explained divergent format preferences. Face-to-face (56%): For older adults living alone (52%), community classes provided vital social contact. Online delivery often failed to replicate camaraderie and peer support [26], reinforcing preference for in-person sessions. Online/no preference (44%): Conversely, 16% favoured tele-exercise and 28% expressed no preference. For them, virtual delivery reduced social obligations and performance pressure, aligning with reports that some older adults prefer minimal socialising during exercise [30,31].

### Motivation

From Scaffolding to Habits (COM□B: Reflective & Automatic; TDF: Beliefs about Consequences, Reinforcement, Habit)

Engagement showed that low education and solitary living need not hinder digital health uptake when material access is addressed. Providing hardware, connectivity, and home onboarding dismantled the digital divide [32]. Sustained adherence followed two pathways:

- **Reflective Motivation:** Reinforced by perceived gains in strength, flexibility, and balance (Theme 3).
- **Automatic Motivation:** Established through routine, scheduled sessions (Theme 3). From a theoretical perspective, these pathways are successful because they satisfy older adults’ core psychological needs. A recent qualitative review framed by Self-Determination Theory found that interventions enhance motivation when they foster competence (e.g., through feedback and mastery), autonomy (e.g., through choice), and relatedness (e.g., through peer support), which aligns directly with the mechanisms observed in our study [33].

Convenience and reduced logistical barriers supported this integration [34,35]. For those preferring online delivery, supervised home-based video programmes offer a sustainable mode of exercise [35]. Tele-exercise thus provides a safe, adaptive alternative when travel is hazardous, provided remote coaching scaffolds both physical and psychological safety.

### Methodological Rigor and Theoretical Pipeline

A primary strength of this study lies in its systematic, theory-driven design pipeline spanning the TDF [36], the COM-B model, the BCW [17], and the BCTTv1 [24]. The 14-domain TDF enabled deductive categorisation of barriers and enablers, capturing nuances such as deficits in self-efficacy, emotional regulation, and environmental resources. Mapping these determinants into COM-B and cross-referencing with BCW functions established a logic model, while appraisal against APEASE (Affordability, Practicability, Effectiveness, Acceptability, Side-effects, and Equity) criteria safeguarded feasibility and equity.

### The Critical Shift: Evaluated Protocol vs. Future Recommendations

The evaluated protocol operationalised Enablement, Training, and Environmental Restructuring through BCTs such as Graded tasks (8.7), Instruction (4.1), and Social support (3.2, 3.3) [37]. Yet qualitative findings revealed limitations in unstandardised home settings, prompting a shift toward participant-centred, structural co-regulation. Recommendations emphasise higher-order BCTs: proactive Action planning (1.4), Restructuring the physical environment (12.1), and Restructuring the social environment (12.2). Evidence shows interventions integrating >3 BCTs are more effective in promoting activity and reducing sedentary behaviour [38]. Sustaining adherence thus depends less on repeated instruction and more on optimising Physical and Social Opportunity [10]. This aligns with evidence demonstrating that tailored, multifaceted implementation strategies are effective for improving healthcare professional adherence to falls prevention guidelines[9]. Integrating these structural BCTs elevates tele-exercise from remote clinical adaptation to a resilient, context-aware complex intervention.

### Strength and Limitations

A major strength of this study is its use of the COM-B model and TDF to explain tele-exercise adoption, translating qualitative insights into actionable BCTs and offering scalable guidance for digital health. It also captures the experiences of a vulnerable group—older adults with high fall risk, low education, and high rates of living alone (52%)—showing that digital exclusion can be mitigated through targeted support.

Limitations include a socio-demographically homogenous sample (single ethnicity, 80% female), restricting generalisability, and high attendance (85.5%) and enjoyment (PACES), which may reflect selection bias. The coupon, though ethically intended as compensation, may have introduced minor volunteer bias and social desirability bias. participants who volunteered and remained engaged were likely more motivated and health-conscious than the broader older adult population. To reduce these risks, participants were assured that compensation was guaranteed regardless of feedback and encouraged to provide critical views.

## Conclusion

This study shows tele-exercise is feasible and acceptable for community-dwelling older adults at risk of falls, challenging assumptions that low digital literacy or socioeconomic barriers preclude digital health adoption. Using the COM-B model and TDF, it demonstrates that technology-related anxiety and physical limitations can be mitigated through targeted BCTs. Mastery of the technology shifted emotions from anxiety to pride, reinforcing digital self-efficacy.

Despite high adherence and enjoyment, over half preferred f2f delivery, indicating that format preference does not dictate engagement. To ensure safety and engagement, future programmes should embed identified BCTs within a structured framework. Addressing structural and psychological barriers can strengthen self-efficacy and sustain remote participation, offering a practical pathway to active ageing.

## Ethics approval and consent to participate

The study conforms to the Declaration of Helsinki. The study involves human participants and was approved by the Ethics Committee of the College of Professional and Continuing Education, The Hong Kong Polytechnic University (RC/ETH/H/0029). All participants provided informed consent to participate in the study prior to their involvement.

## Data availability

The datasets used and analysed during the current study are available from the corresponding author on reasonable request.

## Declaration of generative AI and AI-assisted technologies in the writing process

During the preparation of this work the authors used Copilot and Gemini in order to improve the readability and language of the manuscript. After using this tool/service, the authors reviewed and edited the content as needed and take full responsibility for the content of the published article.

## Acknowledgements

The authors are grateful to the Centre for Ageing and Healthcare Management Research (CAHMR), College of Professional and Continuing Education, The Hong Kong Polytechnic University for coordination with the elderly services providers i.e. Sik Sik Yuen. Thank Sik Sik Yuen for recruiting older people and all the participants for joining this study. The authors would like to express their gratitude to Mr. Au Wing Ho for his exceptional exercise teaching provided to all the participants. Additionally, the authors extend their heartfelt thanks to the student helpers who offered technical support to the participants.

---

**Supplementary material S2:** Consolidated criteria for reporting qualitative studies (COREQ): 32-item checklist (Tong et al., 2007)

| Domain | Section | Item | Guide Question / Description | Check |
|---|---|---|---|---|
| **Domain 1: Research team and reflexivity** | Personal Characteristics | 1. Interviewer/facilitator | Which author(s) conducted the interview or focus group? | Stated, Methodology--> Interview process |
|  |  | 2. Credentials | What were the researcher's credentials? *e.g., PhD, MD* | Stated, Methodology--> Interview process |
|  |  | 3. Occupation | What was their occupation at the time of the study? | Stated, Methodology--> Interview process |
|  |  | 4. Gender | Was the researcher male or female? | Stated, Methodology--> Interview process |
|  |  | 5. Experience and training | What experience or training did the researcher have? | Stated, Methodology--> Interview process |
|  | Relationship with participants | 6. Relationship established | Was a relationship established prior to study commencement? | Stated, Methodology--> Interview process |
|  |  | 7. Participant knowledge of the interviewer | What did the participants know about the researcher? *e.g., personal goals, reasons for doing the research* | Stated, Methodology--> Interview process |
|  |  | 8. Interviewer characteristics | What characteristics were reported about the interviewer/facilitator? *e.g., Bias, assumptions, reasons and interests in the research topic* | Interests in the research topic |
| **Domain 2: Study design** | Theoretical framework | 9. Methodological orientation and theory | What methodological orientation was stated to underpin the study? *e.g., grounded theory, discourse analysis, ethnography, phenomenology, content analysis* | Behaviour Change Wheel – COM-B, TDF, BCTTv1 |
|  | Participant selection | 10. Sampling | How were participants selected? *e.g., purposive, convenience, consecutive, snowball* | Purposive |
|  |  | 11. Method of approach | How were participants approached? *e.g., face-to-face, telephone, mail, email* | Face-to-face or via Zoom |
|  |  | 12. Sample size | How many participants were in the study? | 25 |
|  |  | 13. Non-participation | How many people refused to participate or dropped out? Reasons? | 0 |
|  | Setting | 14. Setting of data collection | Where was the data collected? *e.g., home, clinic, workplace* | Their home or community centre |
|  |  | 15. Presence of non-participants | Was anyone else present besides the participants and researchers? | No |
|  |  | 16. Description of sample | What are the important characteristics of the sample? *e.g., demographic data, date* | Table 1 and 2 |
|  | Data collection | 17. Interview guide | Were questions, prompts, guides provided by the authors? Was it pilot tested? | Yes |
|  |  | 18. Repeat interviews | Were repeat interviews carried out? If yes, how many? | No |
|  |  | 19. Audio/visual recording | Did the research use audio or visual recording to collect the data? | Video recording |
|  |  | 20. Field notes | Were field notes made during and/or after the interview or focus group? | No separate field notes were taken. The sessions were video-recorded, which allowed for the subsequent capture of non-verbal behaviors and verbal transcripts during the analysis stage. |
|  |  | 21. Duration | What was the duration of the interviews or focus group? | Stated, Methodology--> Interview process |
|  |  | 22. Data saturation | Was data saturation discussed? | Yes, Methodology--> Interview process |
|  |  | 23. Transcripts returned | Were transcripts returned to participants for comment and/or correction? | No |
| **Domain 3: Analysis and findings** | Data analysis | 24. Number of data coders | How many data coders coded the data? | Three |
|  |  | 25. Description of the coding tree | Did authors provide a description of the coding tree? | Table 3, Figure 1, Supplementary Table |
|  |  | 26. Derivation of themes | Were themes identified in advance or derived from the data? | Derived from the data |
|  |  | 27. Software | What software, if applicable, was used to manage the data? | NVivo 14 |
|  |  | 28. Participant checking | Did participants provide feedback on the findings? | No |
|  | Reporting | 29. Quotations presented | Were participant quotations presented to illustrate the themes/findings? Was each quotation identified? *e.g., participant number* | Yes |
|  |  | 30. Data and findings consistent | Was there consistency between the data presented and the findings? | Yes |
|  |  | 31. Clarity of major themes | Were major themes clearly presented in the findings? | Yes |
|  |  | 32. Clarity of minor themes | Is there a description of diverse cases or discussion of minor themes? | Yes |

Tong, A., Sainsbury, P., & Craig, J. (2007). Consolidated criteria for reporting qualitative research (COREQ): a 32-item checklist for interviews and focus groups. *International Journal for Quality in Health Care*, *19*(6), 349–357. https://doi.org/10.1093/intqhc/mzm042

**Supplementary material S4:** Mapping of barriers and facilitators to BCW intervention functions and BCTTv1 techniques, with proposed refinements appraised against APEASE.

| Theme | Sub-theme | Intervention Functions (BCW) | Recommended BCTTv1 Techniques | Evaluated Protocol Components (Current Programme) | Proposed Refinements for Future Interventions^, Informed by APEASE |
|---|---|---|---|---|---|
| Theme 1: Technological Skills, Access & Support | Barrier: Technological Limitations Affecting Participation | Environmental restructuring, Enablement, Education | 4.1 Instruction on how to perform the behaviour<br>12.5 Adding objects to the environment | • Hardware Provision: Loaned 4G-enabled tablets, stands, and SIM cards.<br>• Remote Tech Support: Team used remote-access software to troubleshoot tablets prior to class. | • Assess visual needs, available exercise space, and camera coverage during onboarding.<br>• Optimise the supplied device and positioning first;<br>• Offer larger displays or a wide-angle lens attachment or TV casting where compatible, acceptable, and affordable.<br>• Verify that the selected arrangement allows participants to see demonstrations and instructors to observe relevant movements, without requiring additional household equipment. |
|  | Facilitator: Confidence Through Guided Practice | Training, Enablement | 3.2 Social support (practical)<br>6.1 Demonstration of the behaviour<br>8.1 Behavioural practice/rehearsal | • Hands-on Device Onboarding: In-person, step-by-step tablet orientation, charging setup, and guided login drill prior to class 1. | • Standardise the core onboarding content while tailoring its delivery to digital confidence and support needs.<br>• Offer hands-on in-person onboarding where needed, supplemented by accessible pictorial instructions and repeated login practice.<br>• Provide supported remote onboarding for participants able and willing to use it, with access to refresher support. |
| Theme 2: Coach Support & Exercise Adaptability | Barrier: Limited Responsiveness in Online Supervision | Training, Environmental restructuring, Enablement | 2.2 Feedback on behaviour<br>4.1 Instruction on how to perform the behaviour | • Visual Anchoring: Instructor pinned permanently on participant screens.<br>• Large Display Surveillance: Instructor monitored class via a 100-inch screen.<br>• Capped Cohorts: Kept classes small (8–11 participants). | • Provide immediate feedback for safety-critical movement issues during exercise, supplemented by a brief, optional post-class question period.<br>• Offer targeted individual follow-up when correction cannot be delivered adequately within the group session.<br>• Pilot the staffing requirements and assess participants’ perceived adequacy of feedback. |
|  | Facilitator: Supportive Coaching for Effective Participation | Modelling, Enablement, Training | 3.2 Social support (practical)<br>3.3 Social support (emotional)<br>8.7 Graded tasks | • Relatable Role Model: Engaged a peer instructor in their late 60s who synchronously demonstrated all movements.<br>• Dynamic Adaptability: Regressed difficult exercises and allowed flexible exertion (e.g., 60–70% effort). | • Incorporate brief participant choice among instructor-approved exercises or adaptations consistent with programme objectives and individual ability.<br>• Invite suggestions for subsequent sessions, while retaining appropriate progression and core programme content.<br>• Assess whether these choices improve perceived autonomy and engagement. |
| Theme 3: Health Motivation & Goal Achievement | Barrier: Movement challenges linked to health conditions | Training, Enablement, Persuasion | 8.7 Graded tasks<br>15.1 Verbal persuasion about capability | • Safe Baseline (Otago Level A): Standardized starting tier using sturdy chair support to build early confidence.<br>• Verbal Reinforcement: Continuous positive verbal praise and encouragement during exercises. | • Conduct a pre-intervention assessment of health conditions, functional ability, support needs, and the home exercise environment to inform starting levels and adaptations.<br>• Use remote assessment where appropriate; arrange in-person assessment or clinical referral when remote assessment is insufficient or raises safety concerns.<br>• Review exercise levels as participants progress. |
|  | Facilitator: Multidimensional Health Gains and Habitual Routine Fostering Functional Independence | Training, Enablement, Incentivisation | 1.1 Goal setting (behaviour)<br>1.3 Goal setting (outcome)<br>2.7 Feedback on outcome(s) of behaviour<br>8.1 Behavioural practice/rehearsal<br>8.3 Habit formation | • Concrete Target Setting: Set functional milestones (e.g., unsupported single-leg stance near chair).<br>• Fixed Scheduling: Enforced consistent 3x/week timetable to cultivate automatic habit. | • Support routine formation through consistent scheduling, participant-defined functional goals, and simple progress feedback.<br>• Where additional monitoring is justified, pilot optional wearable measures validated for the intended tasks and population, alongside low-technology alternatives.<br>• Evaluate measurement accuracy, participant burden, costs, privacy, and whether monitoring improves engagement or outcomes. |
| Theme 4: Emotional Journey with Technology | Barrier: Initial Fear and Anxiety | Enablement, Training, Persuasion | 11.2 Reduce negative emotions<br>15.1 Verbal persuasion about capability | • Affective Scaffolding: In-home introductory session focused on alleviating tele-conferencing anxiety, reassuring participants against device-handling fears, and establishing emotional trust with the research team. | • Offer optional one-to-one digital buddy support from trained staff, peers, or community volunteers, matched to participants’ language, preferences, and support needs.<br>• Establish supervision, confidentiality, safeguarding, and referral arrangements.<br>• Use guided practice that progressively builds independent device use rather than ongoing reliance on the buddy. |
|  | Facilitator: Pride and Happiness After Mastering Technology | Persuasion, Incentivisation | 10.4 Social reward | • Mastery Commendation: Systematic verbal praise from instructor and team celebrating digital navigation milestones and exercise completion. | • Recognise individually meaningful digital and exercise achievements through specific verbal feedback and, where preferred, simple certificates or digital badges.<br>• Emphasise personal progress rather than competition, and provide equivalent non-digital recognition.<br>• Assess whether participants find the approach motivating and appropriate. |
| Theme 5: Home Environment & Accessibility | Barrier: Constraints of the Home Environment | Environmental restructuring, Enablement | 1.4 Action planning<br>12.1 Restructuring the physical environment<br>12.3 Avoidance/reducing exposure to cues for the behaviour | • Protected Timetable: Fixed class hours scheduled to avoid household meal and family rush times. | • With participant consent, provide brief guidance to household members on reducing avoidable distractions and maintaining a safe exercise area.<br>• Agree realistic arrangements for shared spaces and offer flexible session options where feasible.<br>• Provide equivalent planning support for participants living alone or without household assistance. |
|  | Facilitator: Convenience of Exercising Without Travel | Environmental restructuring, Enablement | 12.1 Restructuring the physical environment<br>12.5 Adding objects to the environment | • Safe Exercise Zone Setup: In-home identification of a well-lit, clutter-free space utilizing a sturdy home chair for balance support. | • Introduce a brief pictorial pre-session checklist covering a clear exercise area, suitable lighting and device positioning, and stable support where required.<br>• Practise its use during onboarding and provide assisted checks when needed.<br>• Specify how participants should contact the instructor and what to do if the setup is unsafe or the connection fails. |
| Theme 6: Social Interaction Dynamics | Barrier: Limited Real-time Chat & Domestic Distractions | Training, Enablement, Environmental restructuring | 2.2 Feedback on behaviour<br>12.1 Restructuring the physical environment<br>12.2 Restructuring the social environment | • Name-Specific Verbal Cueing: Instructor systematically addressed participants by name for real-time form correction and safety check-ins.<br>• Acoustic Management Protocol: Pre-class reminders to turn off household televisions/radios to minimize audio interference while preserving open microphones for safety monitoring. | Communication and safety:<br>• Establish and rehearse a session communication protocol that enables participants to request help promptly.<br>• Select microphone arrangements according to group size, household noise, participant needs, and privacy preferences, rather than applying a universal open-microphone rule.<br>• Include instructor check-ins and a clear response plan for reported difficulties or connection loss.<br>Social interaction:<br>• Offer brief, optional instructor-facilitated social interaction before or after exercise, keeping conversation separate from tasks requiring focused movement.<br>• Start with a simple whole-group format; pilot breakout rooms only where participants can use them comfortably and staffing permits. |
|  | Facilitator: Dual Mechanisms: Offline Community Bonding vs. In-Class Cognitive Motor Focus | Enablement, Environmental restructuring | 3.1 Social support (unspecified)<br>3.3 Social support (emotional) | • Optional Post-Class Chat: Offered 10–15 minutes of open, unstructured socializing post-workout while allowing focused participants to log off immediately. | • Offer an opt-in moderated messaging group for participants seeking contact outside sessions, subject to platform accessibility and privacy preferences.<br>• Establish consent procedures, communication boundaries, and guidance on appropriate content, clarifying that the group is not an emergency service.<br>• Provide non-app alternatives and assess moderation workload, uptake, and perceived social benefit. |

\*BCTs were mapped using BCTTv1 taxonomy and verified against the BCW guidelines.
^Future recommendations represent proposed refinements derived from the qualitative findings and considered against APEASE dimensions. Their effectiveness, resource requirements, unintended consequences, and equity implications require prospective evaluation; inclusion does not imply that all components were tested in the current trial.

